# TrkA abundance is increased in cutaneous nerves in bortezomib-induced neuropathy

**DOI:** 10.1101/2025.07.02.25330620

**Authors:** Yuying Jin, Nadine Cebulla, Daniel Schirmer, Eva Runau, Leon Flamm, Calvin Terhorst, Laura Jähnel, Johanna Güse, Nicola Giordani, Annett Wieser, Julian Brennecke, Annemarie Sodmann, Robert Blum, Claudia Sommer

**Affiliations:** Department of Neurology, University Hospital of Würzburg, Würzburg, Germany

## Abstract

**Background and objectives:** Tropomyosin receptor kinase A (TrkA), a high-affinity receptor for nerve growth factor (NGF), is highly expressed in nociceptive neurons and plays a key role in nociception. Increased NGF levels have been related to local angiogenesis. This study investigates skin biopsies of multiple myeloma patients who developed peripheral neuropathy during bortezomib treatment for localization and abundance of TrkA.

**Methods:** Multiple myeloma patients with bortezomib-induced peripheral neuropathy (BIPN) were recruited at the University Hospital Würzburg between 2021 and 2024. Age-and sex-matched controls without neuropathy were enrolled for comparison. Skin biopsies from the distal leg were analyzed to determine intraepidermal nerve fiber density (IENFD) and area, TrkA mean fluorescence intensity (MFI) and gene expression levels of TrkA and NGF. Additionally, the proximity between nerve fibers and blood vessels was measured.

**Results:** We enrolled 50 patients with BIPN (age 64.1 ± 9.2 years; 76% male) and 27 controls (age 60.7 ± 7.1 years; 81.5% male). Sensory abnormalities included reduced sural nerve action potential amplitude (63.3%), elevated cold (36%) and heat pain detection thresholds (44%) compared to normative population values (95% CI). IENFD was reduced in patients (median 2.6 [1.6–4.2] fibers/mm) compared to controls (4.7 [3.1–6.8] fibers/mm; p < 0.05). TrkA protein abundance in epidermal nerve fibers (normalized MFI) was higher in patients (0.93 [0.56–1.25]) than controls (0.13 [0.08–0.17]; p < 0.0001) and moderately correlated with the number of bortezomib treatment cycles (r = 0.36, p < 0.05). TrkA mRNA levels did not differ between groups, while NGF mRNA levels were lower in BIPN patients (p = 0.01). Among BIPN patients, those reporting pain had higher NGF mRNA levels than those without pain (p = 0.04). Dermal vascular area was increased in BIPN patients (926.0 [714.8–1710] µm^2^) compared to controls (705.1 [496.5–1190] µm^2^; p < 0.05).

**Discussion:** This study identifies reduced IENFD, increased TrkA abundance in remaining fibers, and enhanced dermal vascularization in multiple myeloma patients with BIPN. These findings raise the possibility that dysregulated NGF/TrkA signaling may contribute to small fiber pathology and the associated alterations in sensory function observed in BIPN.

## 1. Introduction

Multiple myeloma (MM) is the second most common hematological malignancy, and bortezomib (BTZ), a first-line treatment for MM, reversibly inhibits the threonine site of the 26S proteasome, leading to tumor cell cycle arrest and apoptosis ^1^. Despite efforts to mitigate the BTZ side effects through combination therapies and dose adjustments ^2-4^, BTZ-induced peripheral neuropathy (BIPN) remains a significant clinical challenge. Among patients experiencing peripheral neuropathy, 13-36% report neuropathic pain, which typically presents in a stocking-and-glove distribution and severely impairs quality of life ^5, 6^. Notably, in some cases, neuropathic pain persists even after discontinuation of BTZ treatment ^6^. The lack of reliable biomarkers for early prognosis and risk stratification hampers timely intervention and personalized treatment.

Neuropathic pain involves fundamental mechanisms in which the tropomyosin receptor kinase A (TrkA), a high-affinity receptor for nerve growth factor (NGF), plays a key role ^7^. In animal models of peripheral neuropathy, elevated protein levels of NGF, TrkA, and phosphorylated TrkA (p-TrkA) have been observed in dorsal root ganglia (DRG) ^8^, with further activation of the p38/MAPK pathway ^9^. Meanwhile, NGF/TrkA signaling enhances the channel activity of transient receptor potential vanilloid 1 (TRPV1) and promotes the release of calcitonin gene-related peptide (CGRP) ^8, 10^ in chemotherapy-induced peripheral neuropathy (CIPN) models, via the PI3K/Akt ^11^ and PKC/NF-kB ^10^ pathways, respectively. Beyond neuronal sensitization, the NGF/TrkA signaling may modulate vascular support mechanisms essential for nerve repair. In CIPN models, impaired vasa nervorum perfusion and density ^12, 13^ correlate with neuropathy, while VEGF therapy mitigates these deficits ^12^. Notably, NGF/TrkA signaling induces VEGF release via the Akt-NO axis ^14^, suggesting a dual role in both pain pathogenesis and vascular regeneration.

Based on these findings, we hypothesized that NGF-TrkA signaling may contribute to BIPN and investigated relevant biomarkers in skin biopsies from patients and controls to assess its potential role.

## 2. Methods

### 2.1. Study Population

The study included 50 patients diagnosed with BIPN, recruited from the University Hospital Würzburg between July 2021 and November 2024. All participants had a confirmed diagnosis of MM based on the International Myeloma Working Group guidelines ^15^ and were either currently undergoing (n = 29) or had previously completed (n = 21) treatment with BTZ.

Peripheral neuropathy was identified through abnormal findings in at least one of the following assessments: sensory or motor function evaluations, quantitative sensory testing (QST), or electrophysiological studies. Among the participants, 19 reported experiencing neuropathic pain as defined by a grading system ^16^.

A control group of 27 healthy individuals was recruited from the local community through online advertisements. These participants were screened to ensure they had no history of neurological disorders or conditions that could affect peripheral nerve function.

The study protocol received approval from the Ethics Committee of the Medical Faculty at the University of Würzburg (#98/20) and adhered to the ethical principles outlined in the Declaration of Helsinki. Written informed consent was obtained from all participants prior to their inclusion. Furthermore, the study was prospectively registered in the German Clinical Trials Register (DRKS00025196), ensuring compliance with the registry’s disclosure requirements before the commencement of the research.

### 2.2. Function of Sensory Nerves

Sensory nerve function was assessed using both electrophysiological studies and QST. Neurophysiological assessments were conducted on the left sural nerve and sensory median nerve to evaluate large fiber function. Sensory nerve action potentials (SNAP) amplitude and sensory nerve conduction velocity (NCV) were recorded following standard methods ^17^.

QST was performed on the dorsum of the right foot of patients following the standardized protocol established by the German Research Network on Neuropathic Pain (DFNS) ^18^. This protocol evaluates a comprehensive range of sensory parameters, including thermal thresholds (cold detection threshold [CDT], warm detection threshold [WDT], thermal sensory limen [TSL]), pain thresholds (cold pain threshold [CPH], heat pain threshold [HPT]), mechanical thresholds (mechanical detection threshold [MDT], mechanical pain threshold [MPT]), sensitivity to mechanical pain stimuli (mechanical pain sensitivity [MPS], wind-up ratio [WUR]), pressure pain threshold (PPT), and vibration detection threshold (VDT).

For the analysis, data for CDT, WDT, TSL, PPT, MPT, MPS, WUR, MDT, and VDT were subjected to log10 transformation. Additionally, to account for potential confounding factors such as age and sex, all data were normalized using z-transformation. The z-score was calculated as follows: z-score = (*X* _single subject_ – Mean _published controls_) / SD _published controls_, referencing published control values ^19^. For ease of interpretation, the algebraic sign of z-score was adjusted so that positive z-score indicated gain-of-function compared to controls, while negative z-score reflected reduced loss-of-function ^20^.

### 2.3. Skin Biopsy and Immunofluorescence Staining

Skin biopsies were obtained from the distal leg of 50 patients and 27 healthy controls using a disposable biopsy punch under local anesthesia with lidocaine. The collected tissue samples were immediately fixed in freshly prepared 4% buffered paraformaldehyde for 30 minutes. Following fixation, the samples were rinsed in 0.1 M phosphate buffer and subsequently transferred to a 10% sucrose solution in phosphate buffer. For preservation, the tissues were snap-frozen in isopentane chilled with liquid nitrogen. The samples were embedded on cork plates using Tissue-Tec® O.C.T. compound (Sakura Finetek GmbH, Freiburg, Germany).

Using a cryostat, sections of 20 µm thickness were prepared. For blocking, sections were incubated in 10% BSA-PBS for 30 minutes, in a humid chamber. The primary antibodies were diluted in a solution of 1% BSA/PBS with 0.3% Triton X-100 (eTable_1) and were applied overnight, at 4°C, in a humid chamber. After washing three times with PBS, sections were incubated with secondary antibodies in 1% BSA/PBS, for 2 hours. After washing with PBS, the samples were embedded in antifade mounting medium containing DAPI (Vector Laboratories, CA, USA) and sealed with CoverGrip. Slides were stored at 4°C.

The sensitivity and specificity of the TrkA and GAP43 antibodies were evaluated (eFigures_1, 2). Detailed methods for primary antibody selection, including sensitivity and specificity assessments, were based on our previous study ^21^. For testing anti-TrkA antibodies, we also used recombinant TrkA expression. For this, Hek293 cells were transfected with a vector (pcDNA3) expressing the complete coding region of TrkA ^22^.

### 2.4. Fluorescence Microscopy

Images (16-bit) were acquired with a Leica DMi8 epifluorescence microscope, equipped with Lumencor’s Bright Engine, and combined with a Thunder Imaging System (Leica Microsystems, Wetzlar, Germany). An HC PL FLUOTAR 20×/0.55 NA objective was used, and images were acquired with a x-y-resolution of 610 nm/pixel and a step size of 1,800 nm. Image post-processing included computational clearing using the Leica LAS X software and subsequent maximum intensity projection.

Confocal images (x, y, z) were acquired at 12-bit using an Olympus FV1000 confocal laser scanning system, a FVD10 SPD spectral detector, and an Olympus UPLSAPO 60×/1.35 NA oil objective, at x-y-resolution of 98 nm/pixel and a step size of 500 nm in z.

### 52.5. Quantitative Analysis

#### 2.5.1. Quantitative Analysis for Fluorescence Intensity

Two fluorescence images per patient were captured along the basement membrane, moving sequentially from left to right to avoid any overlap between fields. Each image included the epidermis, and a downward extension of the epidermis to ensure the inclusion of nerve fibers (eFigure_5A). For unbiased segmentation of the epidermis, dermis, nerves, and blood vessels, a deep learning-based segmentation tool optimized for bioimage analysis was used ^23, 24^, as recently described ^21^. The subepidermis was then extracted by computationally expanding the epidermis by 50 µm and overlapping it with the dermis (eFigure_5B). Quantitative analysis of TrkA and GAP43 labels was restricted to nerve fiber segmentations within ROI.

#### 2.5.2. Quantification of Intraepidermal Nerve Fiber Density (IENFD)

IENFD was initially assessed using a manual counting method ^25^. In this approach, all PGP9.5-immunoreactive fibers crossing the basement membrane in a section were counted (3 sections per patient), and the counts per millimeter length of the tissue section, denoted as IENFD_PGP9.5_. Similarly, nerve fibers labeled with CGRP were denoted as IENFD_CGRP_. The manual count served as the reference for subsequent automated quantification.

For automated quantification, nerve fibers and the epidermis were segmented using a deep learning-based algorithm ^23, 24^, as described above. The intersections between segmented nerve fibers and the epidermal outline were identified to mimic the manual counting process.

#### 2.5.3. Quantification for Vascular-Nerve Fiber Interaction

For investigating blood vessels close to nerve fibers, both structures were segmented using deepflash2. To assess potential interactions, a nerve fiber proximity region was defined as the area within a 5 µm radius surrounding the nerve fibers. Blood vessels located within this region were considered to be in close spatial proximity to the nerve fibers.

### 2.6. RNA Isolation, cDNA Synthesis and Quantitative Real-time PCR

Total RNA was extracted from skin biopsies using the Qiagen miRNeasy Microkit (Cat# 217084, Qiagen, Hilden, Germany). For cDNA synthesis, 250 ng RNA was reverse transcribed in a volume of 100 µl. For qPCR, the following primers were used: TrkA: Hs00176787_m1; NGF: Hs00171458_m1, Thermo Fisher Scientific) and TaqMan probes (Cat# 4444557, Thermo Fisher Scientific, Vilnius, Lithuania). RPL13A primers (Hs04194366_g1) were used as reference housekeeping gene. Reactions were run in MicroAmp Fast 96-well reaction plates (Cat# 4346907, Applied Biosystems, Darmstadt, Germany). Subsequently, 3.5 µl cDNA samples were run in triplicates using a StepOnePlus Real-Time Cycler (Cat# 4376598, Life Technologies, Darmstadt, Germany). The cyclin protocol was: initial incubation at 50°C for 2 minutes, followed by a second incubation at 95°C for 2 minutes, and then 40 cycles of 95°C for 3 seconds and 60°C for 30 seconds. Data evaluation was performed using the 2−ΔΔCT method.

### 2.7. Statistical Analysis

SPSS Statistics 26 software (IBM, Ehningen, Germany) was used to statistical analyze. Shapiro-wilk test was used to test normality of each group. Descriptive statistics of the variables were analyzed and presented as mean with standard deviation (SD, normality distribution) or median with interquartile range (IQR = Q3 -Q1, non-normality distribution). For comparisons between controls and patients, as well as between the painful and painless subgroup of BIPN, the T-test was applied for variables with normal distribution, while the Mann-Whitney U test (a non-parametric test) was used for non-normally distributed variables. Categorical variables, such as sex distribution among patients, were analyzed using the Chi-square (χ^2^) test. Correlations between two continuous variables were assessed using the Spearman rank correlation test.

### 2.8. Data Availability

Anonymized clinical data are available from the corresponding author upon request, pending approval and a signed data-sharing agreement with Würzburg University. Data from patients with BIPN within the KFO5001 project have been partially published previously ^26, 27^. Deep-learning segmentation models, original bioimages, and corresponding predicted segmentations generated by our models are publicly available at Zenodo (DOI: 10.5281/zenodo.15174757).

## 3. Results

### 3.1 Demographic, Neurophysiological, and QST Data of Patients

#### 3.1.1 Demographic and Clinical Characteristics

The study enrolled 50 patients (mean age: 64.1 ± 9.2 years; 76% male), who were divided into two subgroups: 31 patients without pain and 19 patients with pain. As a reference group, 27 healthy volunteers were also included. Demographic and clinical characteristics are summarized in Table 1. Body mass index (BMI) data for the control group were not available.

**Table 1.**
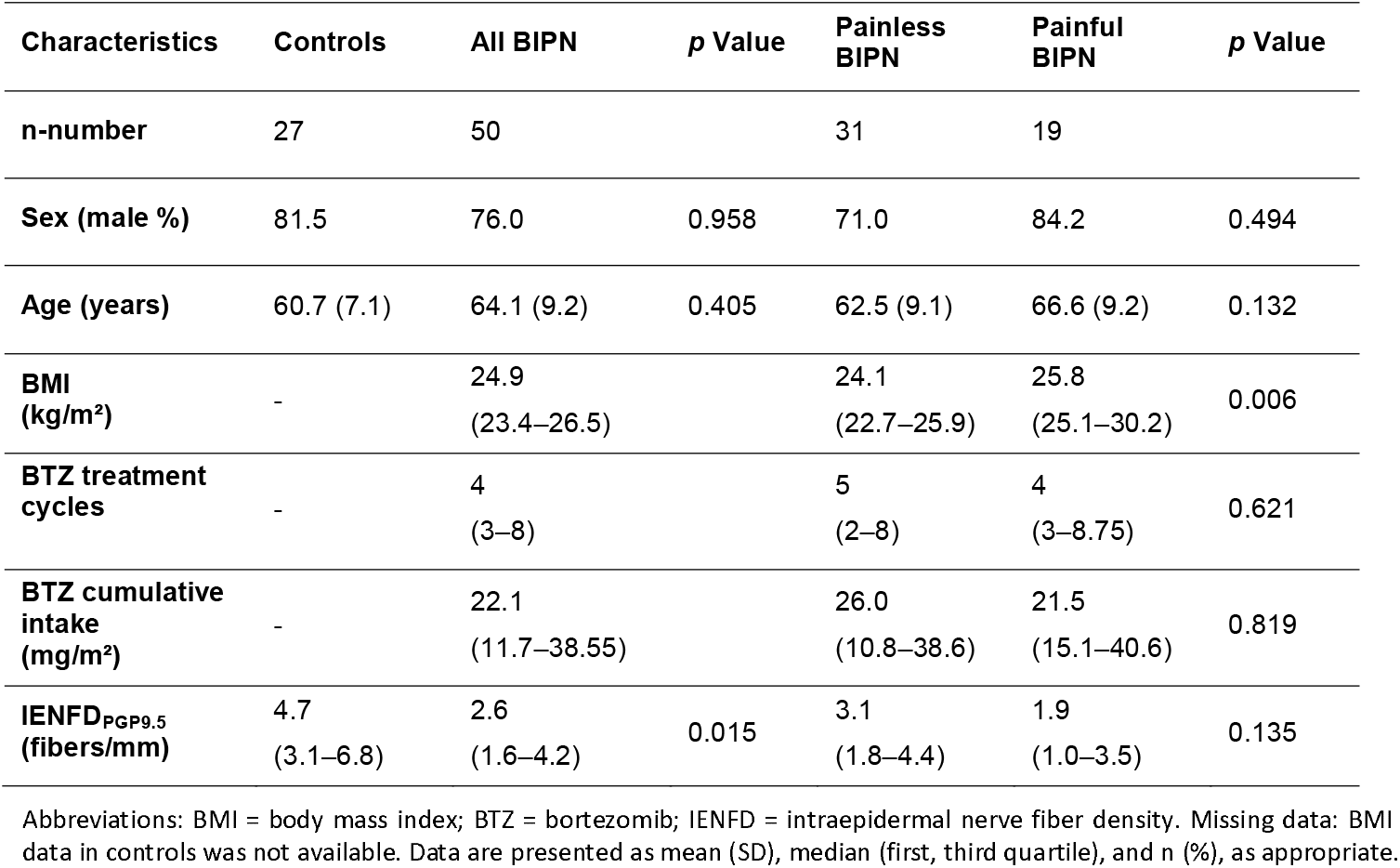
Demographic and Clinical Characteristics of Patients with BIPN.

The numerical rating scale (NRS) based on the maximum rating in the last week without pain medicine, was categorized as follows: no pain (0), n=31, mild (1 - 3), n=4, moderate (4 - 6), n=4, and severe (≥ 7), n=11. For patients who had taken pain medication, the NRS was adjusted to reflect the corrected pain level by incorporating the reported relief from medication (eTable_4).

Statistical analysis revealed no significant differences in age and sex between healthy volunteers and all BIPN patients. Similarly, no significant differences were found between the painful and painless subgroups in terms of age, sex, BTZ treatment cycles, or cumulative dose. However, BMI was significantly higher in the painful subgroup compared to painless subgroup (*p* = 0.017; Table 1). (insert Table 1)

#### 3.1.2 Nerve Conduction Study (NCS) of Sensory Nerves

The nerve conduction parameters (see methods) were evaluated against population normative values (95% confidence intervals, CI); values falling outside this range were considered abnormal.

SNAP amplitudes of the sural nerve were abnormal in 63.3% of evaluable patients with BIPN (31/49; data missing for 1 patient). The painful BIPN subgroup showed a significantly higher rate of impairment (84.2% [16/19]) compared to painless subgroup (50% [15/30]; χ^2^ = 4.48, *p* = 0.034, OR = 5.33, 95% CI: 1.38–19.45) (Figure 1A). The painful subgroup exhibited more severe reductions in sural SNAP amplitudes (median: 3.0 μV, IQR: 0 – 4.5 μV) compared to the painless BIPN (median: 6.65 μV, IQR: 2-12.45 μV) (Figure 1B).

**Figure 1.**
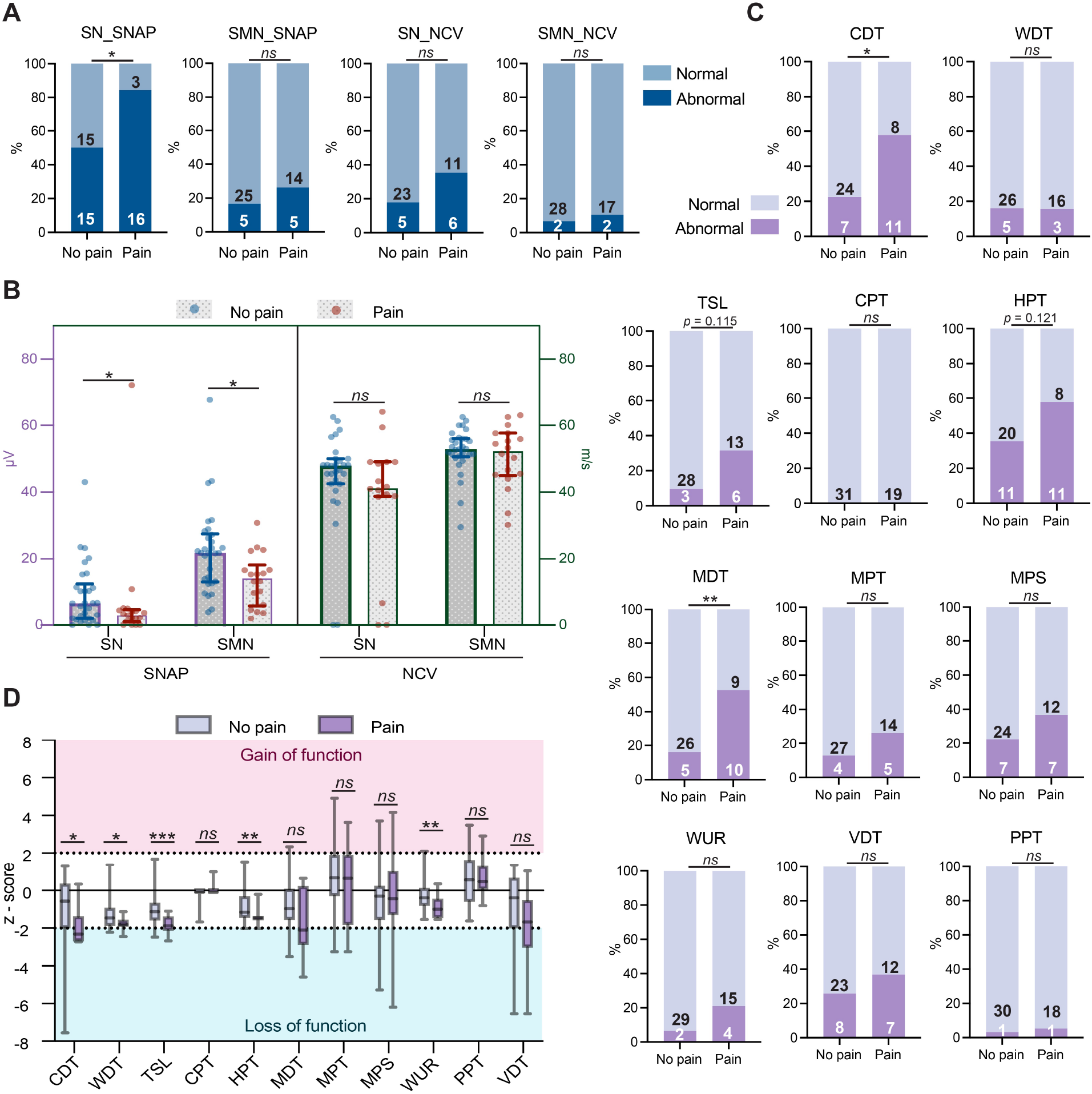
Nerve Conduction Studies (NCS) and Quantitative Sensory Testing (QST) Profiles in Painful and Painless BIPN Subgroups. (A) Percentage of patients with abnormal NCS values (outside 95% CI of normal range). Numbers of patients with normal/abnormal findings are indicated within bars. Group comparisons used Pearson’s χ^2^ test or Yates’s correction, as appropriate. (B) Median values (interquartile ranges) of sensory nerve action potential (SNAP) and nerve conduction velocity (NCV). Missing data: SN_SNAP (n=1), SMN_SNAP (n=1), SN_NCV (n=5), SMN_NCV (n=1). Group comparisons used Mann-Whitney U test. (C) Percentage of patients with abnormal QST values (outside 95% CI of normal range, age- and sex-adjusted). Numbers of patients with normal/abnormal findings are shown within bars. Group comparisons used χ^2^ test, Yates’s correction, or Fisher’s exact test, depending on sample size assumptions. (D) QST z-scores (median ± interquartile range). Positive z-scores indicate gain of function; negative scores indicate loss of function. Mann-Whitney U test was used for group comparisons. Abbreviations: SN = sural nerve; SMN = sensory median nerve; SNAP = sensory nerve action potential; NCV = nerve conduction velocity; CDT = cold detection threshold; WDT = warm detection threshold; TSL = thermal sensory limen; HPT = heat pain threshold; CPT = cold pain threshold; MDT = mechanical detection threshold; MPS = mechanical pain sensitivity; MPT = mechanical pain threshold; VDT = vibration detection threshold; WUR = wind-up ratio; PPT = pressure pain threshold. * *p* < 0.05, ** *p* < 0.01, *** *p* < 0.001, ns: not significant.

Median nerve SNAP amplitudes were abnormal in 20.48% of the BIPN patients (10/49; 1 missing), with abnormal values observed in 26.3% [5/19] of the painful subgroup and 16.7% [5/30] in the painless subgroup (not significant, n.s.) (Figure 1A). However, the painful subgroup showed more pronounced reductions SNAP amplitudes (median: 15.3 μV, IQR: 6.2–18.3 μV) compared to painless subgroup (median: 21.8 μV, IQR: 13.1–27.5 μV) (Figure 1B).

Abnormal nerve conduction velocity (NCV) was observed in 24.4% of evaluable patients for the sural nerve (11/45; 5 missing) with rates of (29.4% [5/17] in the painful subgroup and 21.4% [6/28]) in the painless subgroup.

For the median nerve, abnormal NCV was found in 8.2% of patients (4/49; 1 missing) including 10.5% [2/19] in the painful subgroup and 6.7% [2/30] in painless subgroup. No significant differences were found between subgroups for either nerve (Figure 1A). Similarly, absolute NCV measurements did not differ significantly between subgroups for either the sural or median nerve (Figure 1B).

#### 3.1.3 Quantitative Sensory Testing (QST)

QST parameters were evaluated against population normative values (95% CI) stratified by age and sex ^19^, with values outside this range considered abnormal (including sensitivity and insensitivity).

Among BIPN patients, abnormal rates for QST parameters were as follows: CDT (36% [18/50]), WDT (16% [8/50]), TSL (18% [9/50]), CPT (0% [0/50]), HPT (44% [22/50]), MDT (30% [15/50]), MPT (18% [9/50]), MPS (28% [14/50]), WUR (12% [6/50]), PPT (30% [15/50]), and VDT (4% [2/50]) (Figure 1C). Patients with painful BIPN exhibited significantly higher abnormal values than BIPN patients of the painless subgroup with respect to CDT (57.9% [11/19] in comparison to 22.6% [7/31], *p* <0.05) and MDT (52.6% [10/19] in comparison to 16.1% [5/31], *p <*0.05) (Figure 1C).

Z-score profiles revealed greater loss of function in painful BIPN (vs. painless) in CDT (*p* = 0.018; −2.31 [−2.7 to −1.48] vs. −0.56 [−1.84 to 0.16]), WDT (*p* = 0.038; −1.81 [−1.91 to −1.57] vs. −1.46 [−1.89 to −0.93]), TSL (*p* < 0.001; −1.88 [−2.11 to −1.52] vs. −1.13 [−1.56 to −0.75]), HPT (*p* = 0.008; −1.46 [−1.52 to −1.38] vs. −1.15 [−1.46 to −0.40]), and WUR (*p* = 0.005; −0.99 [−1.47 to −0.63] vs. −0.39 [−0.81 to 0.13]) (Figure 1D).

These findings suggest that BTZ predominantly affects small nerve fibers (C fibers and Aδ fibers) rather than medium or large nerve fibers (Aβ fiber) in patients with multiple myeloma.

### 3.2. Intraepidermal Nerve Fiber Density in Patients with BIPN

Given that electrophysiological data indicated a small nerve fiber phenotype, we investigated whether intraepidermal nerve fiber density (IENFD) is altered in patients with BIPN.

To quantify IENFD, we employed deep learning (DL)-based, nerve fiber annotation using anti-PGP9.5 immunolabeling (IENFD_PGP9.5_) in bioimages of 20-µm skin sections. To compare this with the standardized manual annotation method ^25^, which is typically conducted on 50-µm sections, we assessed the correlation between automated and heuristic nerve fiber counts. DL-based counts showed a significant positive correlation with manual counts (r = 0.351, 95% CI: 0.121–0.545, p < 0.01). As anticipated, automated counts from 20-µm sections were lower than manual counts from 50-µm sections, likely due to differences in section thickness. In the same set of experiments, we also performed DL-model based annotation of CGRP+ nerve fibers (Figure 2A).

**Figure 2.**
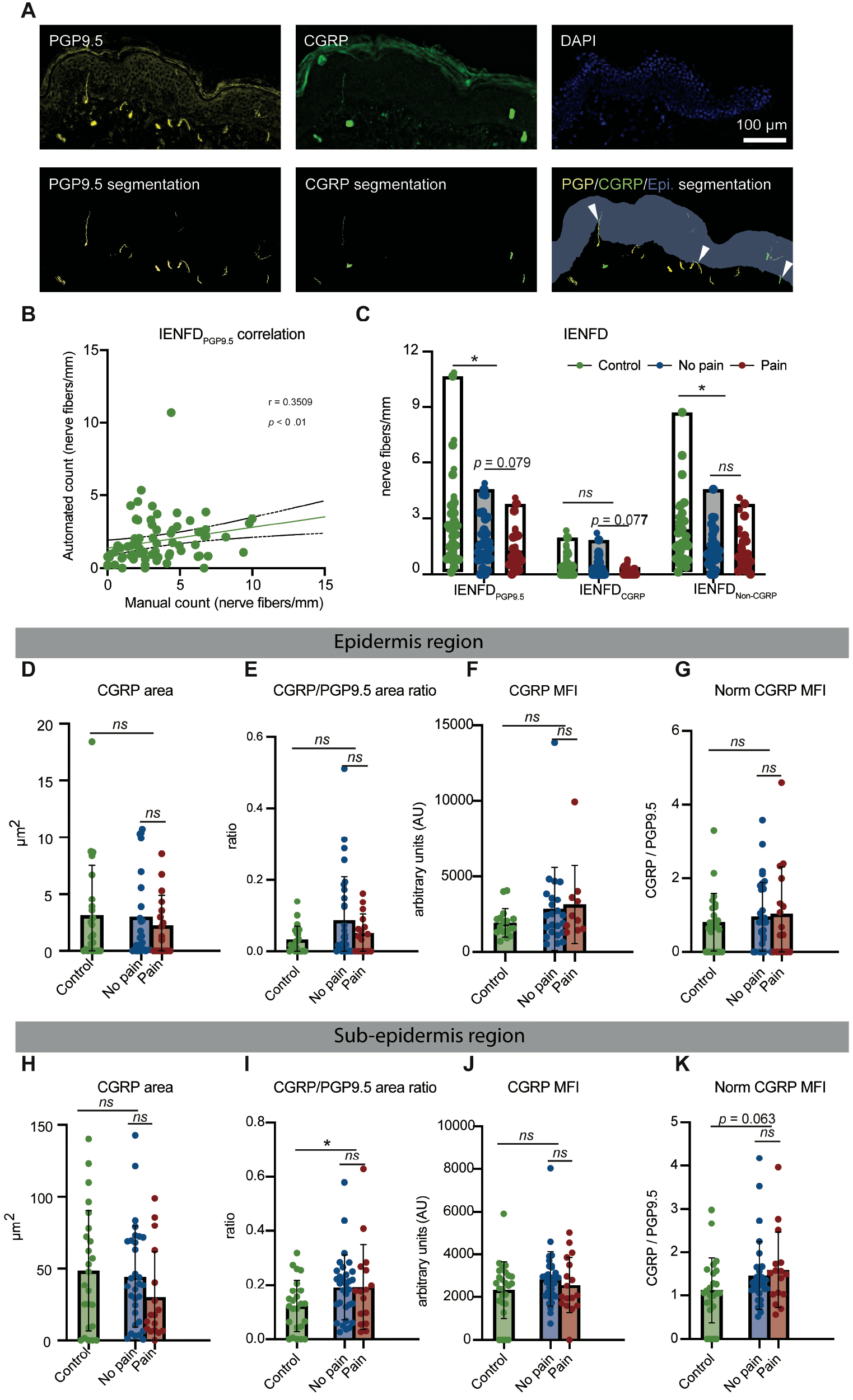
Automated Quantification on PGP9.5 Immunoreactive and Peptidergic Nerve Fibers. (A) Representative images of nerve fibers marked with PGP9.5 (yellow), peptidergic nerve fibers marked with CGRP (green), and nuclei marked with DAPI (blue), alongside their corresponding automated segmentations. (B) Correlation analysis between manual and automated counts of IENFD_PGP9.5_. (C) Automated quantification of IENFD for different types of nerve fibers, including all nerve fibers (PGP9.5+), peptidergic nerve fibers (CGRP+), and non-peptidergic nerve fibers. (D) CGRP+ nerve fiber area in epidermis. (E) Ratio of CGRP+ to PGP9.5+ nerve fiber area in epidermis (range: 0-1). (F) CGRP mean fluorescence intensity (MFI) in epidermis. (G) Normalized CGRP MFI (to PGP9.5 MFI) in epidermis. (H) CGRP+ nerve fiber area in sub-epidermis. (I) Ratio of CGRP+ to PGP9.5+ nerve fiber area in sub-epidermis (range: 0 - 1). (J) CGRP MFI in sub-epidermis. (K) Normalized CGRP MFI (to PGP9.5 MFI) in sub-epidermis. Statistical analysis was performed using correlation analysis for automated and manual count for PGP9.5+ nerve fibers and the Mann-Whitney U test for comparisons between control and patient groups, as well as between painful and painless subgroups. *P <0.05, **P <0.01, ***P <0.001, ns: not significant. Scale bar = 100 µm.

In BIPN patients, we found a decrease in IENFD_PGP9.5_ compared to controls. When the IENFD was analyzed based on anti-CRGP immunofluorescence labelling, IENFD was not different between BIPN patients and controls. It has to be noted that CGRP-immunoreactive fibers were very rare in the epidermis (Figure 2C, Table 2).

**Table 2.**
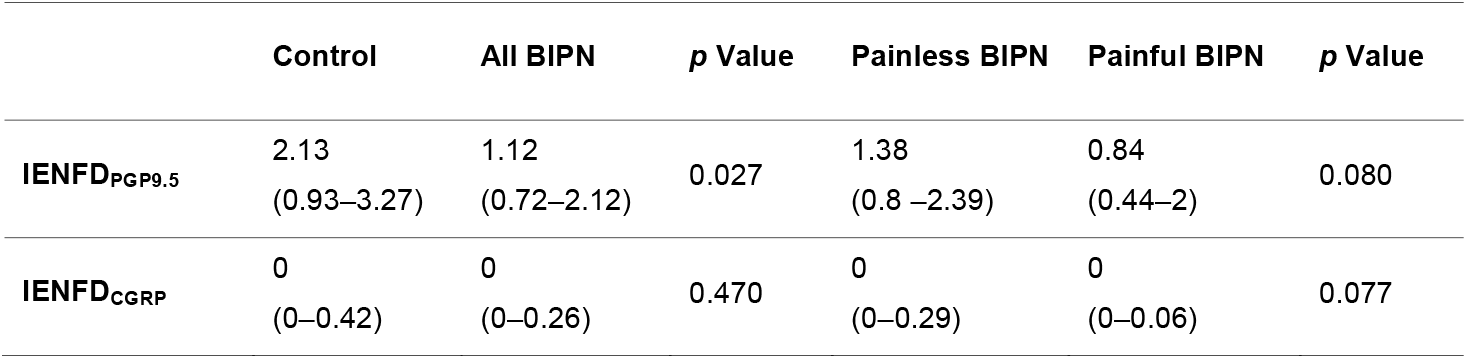
Automated IENFD counts for all nerves, peptidergic nerves, non-peptidergic nerves.

### 3.3. CGRP Immunoreactivity in Human Skin of Patients with BIPN

In the epidermis, the total area occupied by CGRP+ nerve fibers was small and did not differ significantly among controls, and patients with painless or painful BIPN (Figure 2D). The proportion of CGRP^+^ area relative to the total nerve fiber area (PGP9.5^+^) was similarly low in both groups (controls: 2.3%; BIPN: 3.6%) (Figure 2E).

CGRP immunofluorescence intensity (median fluorescence intensity, MFI) was not significantly different between controls and BIPN patients (controls: median 1658, IQR 1314–2195; BIPN: median 2386, IQR 1372–3593; p = 0.199) (Figure 2F).

Likewise, normalized CGRP MFI—calculated as the ratio of CGRP to PGP9.5 MFI—did not differ significantly between groups (controls: median 0.755, IQR 0–1.205; BIPN: median 0.787, IQR 0–1.715; p = 0.199) (Figure 2G).

In the sub-epidermis, the CGRP+ nerve fiber area was 40-fold higher compared to the epidermis, but was not different between controls and BIPN patients (Figure 2H). However, the proportion of the CGRP+ area to total nerve fiber area was 1.5-times higher in BIPN compared to controls (18.3% vs. 12.6%, *p* = 0.027) (Figure 2I). CGRP immunofluorescence intensity values did not differ between controls and BIPN patients (median: 2583, IQR: 1722 – 3058 vs. median: 2685, IQR: 1954 – 3442, *p* = 0.370) (Figure 2J). Moreover, when normalized to PGP9.5 MFI, the normalized CGRP MFI was not significantly different between BIPN patients and controls (Figure 2K).

### 3.4. TrkA Gene Expression and Immunoreactivity in Human Skin

Next we asked whether BIPN is associated with altered expression and abundance of NGF and its receptor TrkA. On the RNA level, qPCR revealed a trend toward lower TrkA expression in the skin of BIPN patients compared to controls (median: 1.349 vs 2.046; Mann-Whitney U=372, *p* = 0.174), with a small effect size (rank-biserial r = -0.21) (Figure 3A). Moreover, NGF RNA expression was significantly decreased in BIPN patients compared to healthy controls (median: 0.92, IQR: 0.69 – 1.14 VS. median: 1.22, IQR: 1.03 – 1.43, *p* = 0.009) (Figure 3A). Within the BIPN cohort, patients with painful neuropathy exhibited higher NGF mRNA levels than those with painless BIPN (median: 1.12, IQR: 0.90–1.14 vs. median: 0.794, IQR: 0.64–1.03, *p* = 0.036) (Figure 3A).

**Figure 3.**
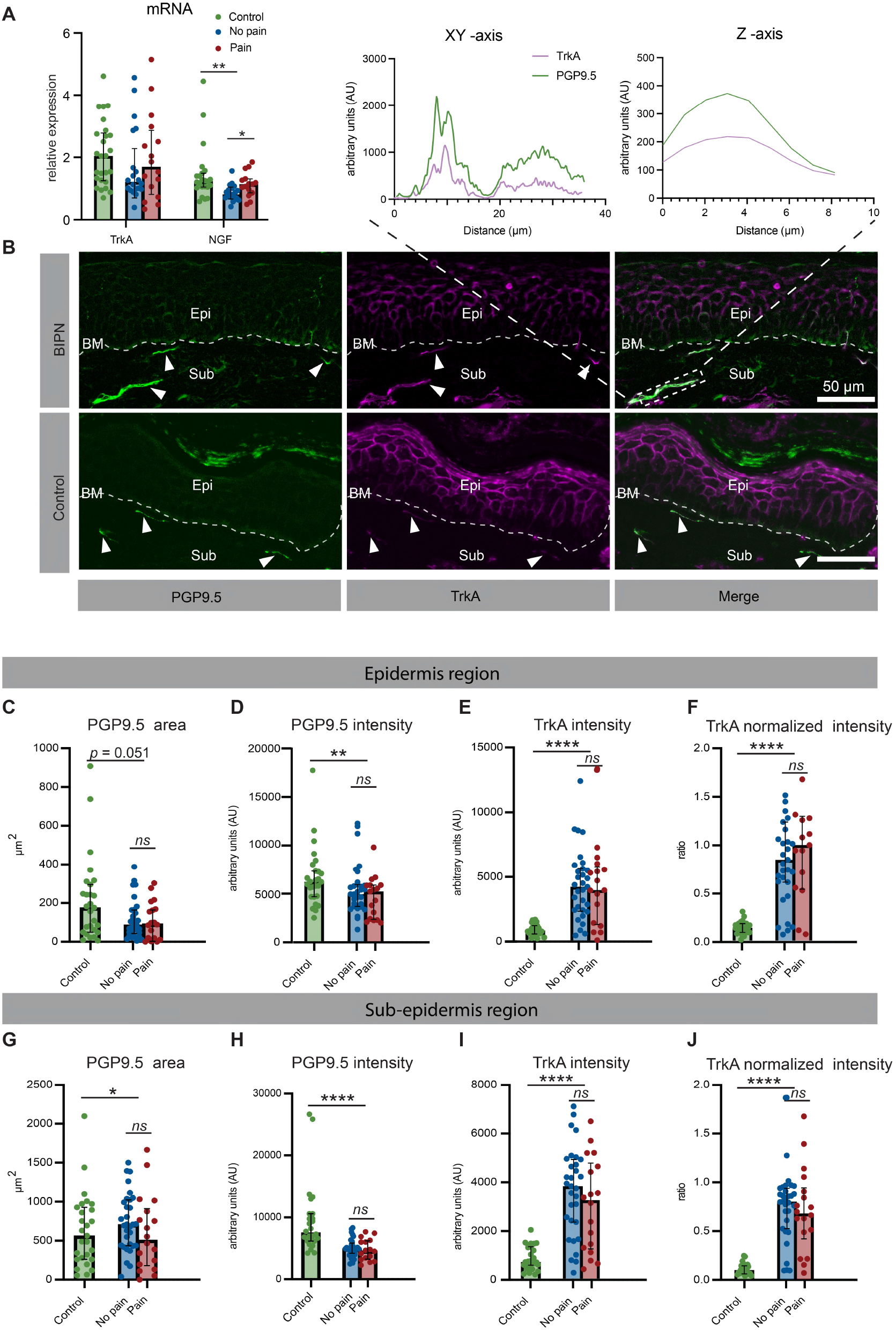
TrkA and NGF in Human Skin. (A) mRNA expression levels of TrkA and NGF in skin biopsy. (B) Top: Colocalization plot for GAP43 and PGP9.5. Bottom: Representative immunofluorescence images showing TrkA expression in control and BIPN, with PGP9.5 used as a reference marker for nerve fibers. (C) Quantification of nerve fiber area (PGP9.5+) in the epidermis region. (D) Mean fluorescence intensity of PGP9.5 in the epidermal region. (E) Mean fluorescence intensity of TrkA in different groups within the epidermal region. (F) TrkA intensity normalized to PGP9.5 in the epidermal region. (G) Quantification of nerve fiber area (PGP9.5+) in the sub-epidermis region. (H) Mean fluorescence intensity of PGP9.5 in the subepidermal region. (I) Mean fluorescence intensity of TrkA in different groups within the subepidermal region. (J) TrkA intensity normalized to PGP9.5 in the subepidermal region. Statistical analysis was performed using the Mann-Whitney U test for comparisons between control and patient groups, as well as between pain and painless subgroups. Scale bar = 50 µm. Abbreviations: BM = basement membrane; Sub = sub-epidermis region; Epi = epidermis region. \**p* < 0.05, ** *p* < 0.01, *** *p* < 0.001, ns: not significant.

At the protein level, anti-TrkA labels showed substantial colocalization with anti-PGP9.5 labels (Figure 3B). In this dataset, quantification of PGP9.5 immunofluorescence revealed a two-fold reduction in total intraepidermal nerve fiber area in BIPN patients compared to controls (Figure 3C). Similarly, PGP9.5 mean fluorescence intensity (MFI) was decreased in BIPN patients (Figure 3D). In contrast, TrkA MFI was more than five times higher in BIPN patients than in healthy controls (Figure 3E). When normalized to PGP9.5, TrkA MFI was elevated by more than six-fold in BIPN patients (Figure 3F). Both absolute and normalized TrkA MFI showed moderate positive correlations with the number of bortezomib (BTZ) treatment cycles (r = 0.3254, *p* = 0.029; r = 0.3637, *p* = 0.014), but neither correlated with cumulative BTZ dose.

The same analysis was applied to nerve fibers in the sub-epidermal region. Here, the PGP9.5 area did not differ significantly between controls and BIPN patients (Figure 3G) although PGP9.5 MFI was decreased in BIPN patients (Figure 3H). In contrast, both TrkA MFI and normalized TrkA MFI were significantly increased—by approximately six-fold—in BIPN patients compared to controls (eTable_2; Figure 3I, J). However, in contrast to the epidermal findings, neither TrkA MFI nor normalized TrkA MFI in the subepidermal region correlated with the number of BTZ treatment cycles or cumulative BTZ dose (*p* > 0.05 for all comparisons).

### 3.5. Vascular-Nerve Fiber Interaction

Given reported effects of NGF-TrkA signaling on blood vessel formation ^28^, we assessed dermal blood vessel area and vessel innervation in the skin samples. The vascular area was increased in BIPN patients compared to controls (median: 926.0, IQR: 714.8 – 1710 vs. median: 705.1, IQR: 496.5 – 1190, *p* = 0.036), with no differences observed between the BIPN subgroups (Figure 4A).

**Figure 4.**
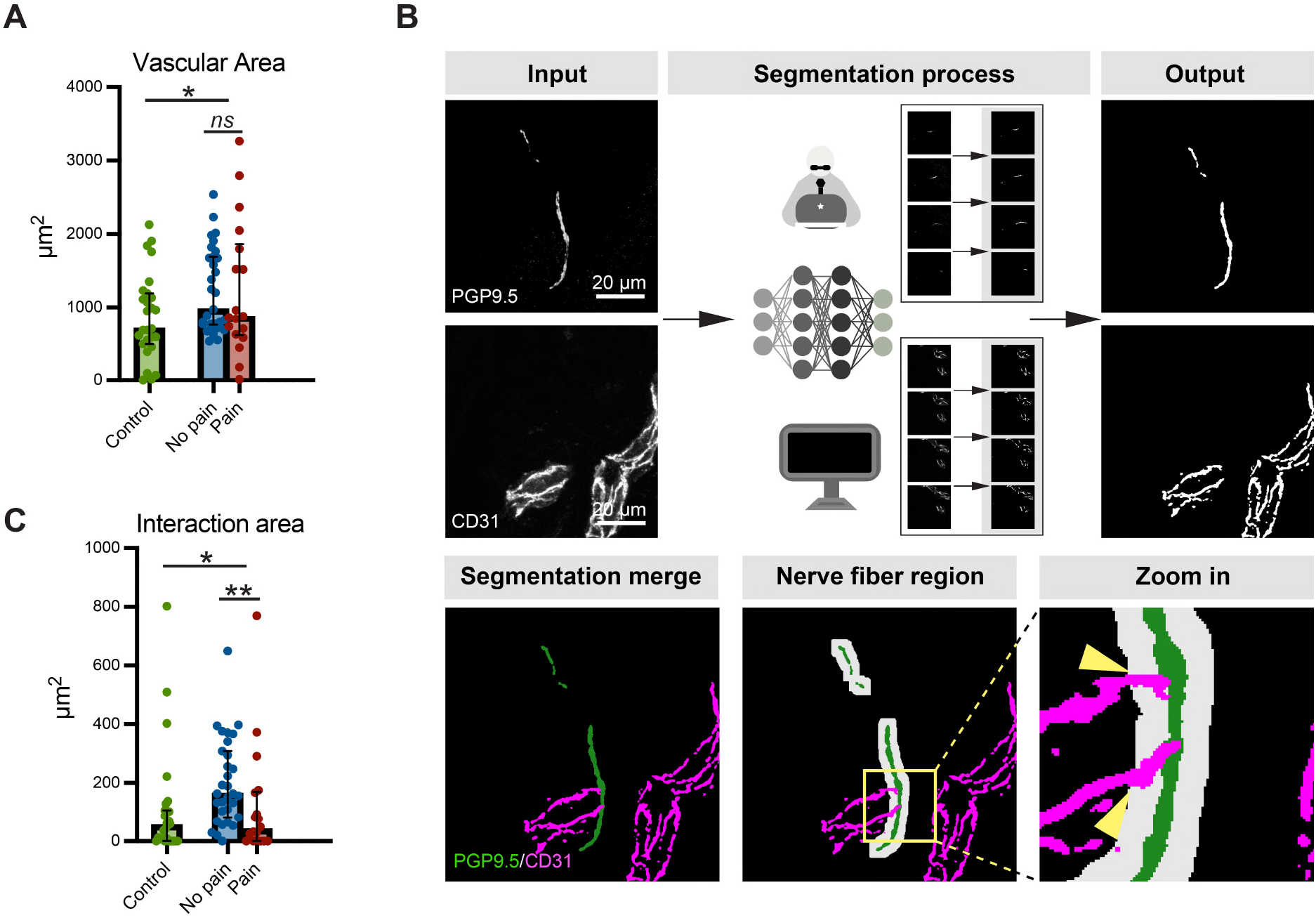
Vascular-Nerve Fiber Interaction in the Dermis Region. (A) Vascular area in the dermal region of control and patient groups, as well as pain and painless subgroups. (B) Workflow for quantifying the interaction area between blood vessels and nerve fibers. Following segmentation of blood vessels and nerve fibers, the interaction area was determined by analyzing the overlap between broadened nerve fibers and blood vessels. (C) Interaction area between blood vessels and nerve fibers in the dermal region in control and patient groups, as well as in pain and painless subgroups. Nerve fiber region was defined as the area within a 5 µm radius surrounding the nerve fibers. Statistical analysis was performed using the Mann-Whitney U test for comparisons between control and patient groups, as well as between pain and painless subgroups. * *p* < 0.05, ** *p* < 0.01, *** *p* < 0.001, ns: not significant. Scale bar = 20 µm.

We also quantified the area of blood vessels located close to the nerve fiber region, defined as the area within a 5 µm radius around the nerve fiber segmentation (see workflow in Figure 4B). In BIPN patients, the vessel-nerve area (blood vessel area within the extended nerve fiber region) revealed two phenotypes:

1. In the painless BIPN subgroup, the area was three times larger than in controls (median: 164.0, IQR: 81.86 – 308.4 vs. median: 54.2, IQR: 0.31 – 105.9, *p <* 0.001) (Figure 4C).
2. The painful BIPN subgroup showed a vessel-nerve area similar to that of controls (median: 48.96, IQR: 0.62 – 167.5) (Figure 4C).

### 3.6. GAP43 Expression and Nerve Regeneration

NGF-TrkA activation has been shown to increase GAP43 immunoreactivity, associated with painful diabetic peripheral neuropathy ^29^. Given the increased TrkA levels, we investigated whether GAP43 immunoreactivity was also increased in case of BIPN.

Immunolabeling showed that GAP43+ nerve fibers largely overlapped with PGP9.5+ nerve fibers in all samples (eFigure_3A). Quantitative analysis revealed no significant differences in GAP43 mean fluorescence intensity (MFI) or normalized GAP43 MFI (relative to PGP9.5) between controls and BIPN patients in either the epidermis or sub-epidermis (eFigure_3B). To explore whether peptidergic fibers might be specifically affected, we also analyzed GAP43 immunoreactivity in CGRP+ nerve fibers. Similarly, GAP43 MFI and normalized GAP43 MFI (relative to CGRP) showed no significant differences between controls and BIPN patients in either the epidermis or sub-epidermis (eFigure_3B, eTable_3). Notably, a decrease in GAP43 MFI was observed on all nerve fibers in the painful subgroup of the BIPN cohort, when compared with the painless subgroup (*p* = 0.041) (eFigure_3B), although no significant differences in normalized GAP43 intensities were observed (eFigure_3B).

## 4. Discussion

In this study, we found decreased intraepidermal nerve fiber density, but increased TrkA abundance on remaining nerve fibers in lower leg skin from patients with BIPN, accompanied by enhanced dermal vascularization. Elevated NGF mRNA levels and increased TrkA immunoreactivity in the painful subgroup suggest that NGF–TrkA signaling may contribute to symptom development in these patients.

Consistent with the reduced IENFD, BIPN patients in our cohort exhibited decreased sensory nerve conduction velocities and reduced sensory nerve action potential amplitudes in the sural nerve, along with thermal sensory deficits – findings that typically accompany a reduction in IENFD_PGP9.5_ ^30, 31^. These results indicate that both large and small sensory nerve fibers are impaired in patients with BIPN.

Notably, axonal degeneration, as assessed by reductions in IENFD and nerve fiber area, did not show a preferential effect on CGRP+ nerve fibers. However, the relative percentage of CGRP+ nerve fiber area (to PGP9.5+ nerve fiber area) and normalized CGRP MFI were increased in the sub-epidermal region of BIPN patients. This finding aligns with earlier studies demonstrating an increase in CGRP+ neurons in the dorsal root ganglion (DRG) of BIPN mouse models ^32^ and higher CGRP+ fiber density in painful diabetic polyneuropathy patients ^33^, suggesting a potential compensatory or maladaptive upregulation of CGRP expression in response to nerve injury.

NGF binding to its receptor TrkA initiates receptor internalization, followed by either recycling to the plasma membrane or degradation via the lysosomal pathway—a process regulated in part by proteasome-mediated deubiquitination ^34^. In the context of BIPN, the observed increase in TrkA immunoreactivity on nerve fibers, despite decreased NGF mRNA expression in whole skin biopsies, may be attributed to the effects of BTZ treatment. As a proteasome inhibitor, BTZ disrupts the ubiquitin-proteasome system, which is responsible for the degradation of approximately 80% of cellular proteins ^35^. Inhibiting proteasomal activity likely interferes with TrkA deubiquitination and degradation, resulting in its accumulation within nerve fibers. Additionally, BTZ-induced toxicity may also impair keratinocytes, a major source of NGF in the skin, leading to reduced NGF mRNA expression in BIPN patients ^36^.

Together, these mechanisms provide a plausible explanation for the increased TrkA immunoreactivity observed in BIPN: impaired proteasomal degradation causes TrkA accumulation, while reduced NGF expression reflects keratinocyte damage. This interpretation is further supported by the moderate positive correlation between the number of BTZ treatment cycles and normalized TrkA MFI.

Experimental studies have shown that NGF injection induces long-lasting thermal hyperalgesia in both animals ^37^ and humans ^38^. Conversely, neutralizing antibodies against NGF or TrkA as well as TrkA mutations, attenuate behavioral responses to noxious stimuli in animal models ^39, 40^. In line with our findings in BIPN, NGF mRNA levels have been reported to be decreased in the skin of patients with peripheral neuropathy (PNP) ^41^. Moreover, clinical phase II trials investigating anti-NGF antibodies for the treatment of diabetic peripheral neuropathic pain and postherpetic neuralgia (PHN) have demonstrated significant pain reduction ^42, 43^.

In CIPN models, reduced nerve blood perfusion and diminished density of vasa nervorum have been reported ^12, 13^. Consistent with these findings, we observed a reduction in blood vessel area within the nerve fiber region in patients with BIPN, despite an overall increase in total dermal blood vessel area. This suggests that the enhanced dermal vascularization in painful BIPN may not confer functional benefit, as these patients continue to experience significant neuropathic symptoms.

In contrast, painless BIPN patients exhibited a different vascular pattern, implying that not just the quantity, but the spatial distribution and functional interaction between blood vessels and nerve fibers may be critical for symptom modulation.

Effective oxygen, nutrient, and hormone exchange likely depends on the close anatomical and functional relationship between nerve fibers and blood vessels ^44^, as observed in the painless BIPN group (eFigure_4).

The usability of GAP43 as a biomarker for peripheral neuropathy remains controversial. While an increased IENFD ratio of GAP43/PGP9.5 was proposed as biomarker to distinguish painful diabetic neuropathic (DNP) from painless DNP ^29, 45^, other studies reported conflicting results. For instance, IENFD of GAP43 was decreased in painful DNP ^46^, and the GAP43/PGP9.5 ratio did not significantly change before and after chemotherapy in patients ^47^. These findings are similar to observations in nerve compression cases before and after surgery ^48^ as well as to our results, further complicating the interpretation of GAP43.

Our research has limitations: First, the method of calculating non-peptidergic nerve fibers by subtracting peptidergic nerve fibers from the total nerve fiber count is overly simplistic. This approach does not account for the substantial overlap in the expression of non-peptidergic and peptidergic markers in human DRG neurons ^49, 50^. Second, skin sections were quantified at a single time point, which limits the ability to capture dynamic changes on nerve fibers. Third, the quantification of bioimages was conducted using a local view (region of interest), which may overlook broader spatial patterns and lead to incomplete or biased data interpretation. Finally, the TrkA antibody used in this study targets total TrkA rather than activated (phosphorylated) TrkA. To further validate TrkA activation, future studies should employ antibodies specific to phosphorylated TrkA or assess downstream signaling pathways, such as PI3K/Akt and MAPK cascades, which are critical mediators of TrkA signaling.

In conclusion, this study suggests that BTZ—a first-in-class proteasome inhibitor used in the treatment of multiple myeloma—damages small nerve fibers and may contribute to pain symptoms in BIPN by impairing the degradation of functional TrkA receptors, leading to their accumulation in remaining nerve fibers.

## Data Availability

Anonymized clinical data are available from the corresponding author upon request, pending approval and a signed data-sharing agreement with Wuerzburg University.

## Author Contributions

Y. Jin: Drafting/revision of the manuscript for content, including medical writing for content; study concept or design; development and implementation of methodology/software for data analysis. C. Sommer: Drafting/revision of the manuscript for content, including medical writing for content; study concept or design; supervision; resources; funding acquisition. N. Cebulla: Major role in the acquisition of clinical data. D. Schirmer: Major role in the acquisition of clinical data. E. Runau: Major role in the acquisition of clinical data. L. Flamm: Major role in the acquisition of clinical data. C. Terhorst: Major role in the acquisition of clinical data. L. Jähnel: Major role in the acquisition of clinical data. J. Güse: Major role in the acquisition of clinical data. N. Giordani: Major role in the acquisition of clinical data. A. Wieser: Major role in the acquisition of clinical data. J. Brennecke: Screening antibodies. R. Blum: Screening antibodies; revision of the manuscript for content; funding acquisition. A. Sodmann: Development and implementation of methodology/software for data analysis.

## Acknowledgements

The authors would like to express their gratitude to Kathleen Stahl for processing skin biopsies and all patients as well as controls for participating in this study.

## Study Funding

This study was conducted as part of the ResolvePAIN project (Project ID: 426503586), which is funded by the Deutsche Forschungsgemeinschaft (DFG, German Research Foundation). Yuying Jin received a scholarship from the Chinese Scholarship Council (CSC, No. 202106230110) to support her participation in this research.

## Disclosure

This study was conducted as part of the ResolvePAIN project KFO5001 (Project ID: 426503586), which is funded by the Deutsche Forschungsgemeinschaft (DFG, German Research Foundation, grants to C. Sommer and R. Blum). Y. Jin received a scholarship from the Chinese Scholarship Council (CSC) to support her participation in this research. The other authors report no relevant disclosures.

## Notes

### Competing Interest Statement

The authors have declared no competing interest.

### Clinical Trial

DRKS00025196

### Author Declarations

Ethics Committee of the Medical Faculty at the University of Wuerzburg (#98/20) gave ethical approval for this work.

