## supplemental figures and tables for "TrkA abundance is increased in cutaneous nerves in bortezomib-induced neuropathy": Supplemental Materials.pdf

### eTable_1

#### Antibody Information

| Target | Host | Clonality | Cat# | RRID | Dilution |
|---|---|---|---|---|---|
| **TrkA, Human** | Goat | polyclonal | AF175 | AB_354970 | 1:500 |
| **CD31, Human** | Mouse | monoclona l | 550389 | AB_2252087 | 1:500 |
| **PGP9.5, Human** | Chicken | polyclonal | NB110-58872 | AB_877619 | 1:200 |
| **GAP43, Human** | Rabbit | polyclonal | ab12274 | AB_2247459 | 1:200 |
| **CGRP, Human** | Mouse | monoclona l | sc-57053 | AB_2259462 | 1:200 |
| **IgG, Goat (Sab to TrkA)** | Donkey | polyclonal | 705-165-147 | AB_2307351 | 1:300 |
| **IgG, Mouse (Sab to CD31/CGRP)** | Donkey | polyclonal | 715-605-150 | AB_2340862 | 1:300 |
| **IgG, Rabbit (Sab to GAP43)** | Donkey | polyclonal | 711-165-152 | AB_2307443 | 1:300 |
| **IgG, Chicken (Sab to PGP9.5)** | Donkey | polyclonal | 703-545-155 | AB_2340375 | 1:300 |

Abbreviations: Sab = secondary antibody; RRID = research resource identifier

### eTable_2

**TrkA/PGP9.5 Immunofluorescence Intensity Quantification**

| | Parameters | Control | All BIPN | *p* Value | Painless BIPN | Painful BIPN | *p* Value |
|---|---|---|---|---|---|---|---|
| Epi | **PGP9.5 area** | 176.1 (49.17–295.4) | 87.95 (28.82–166.1) | **0.051** | 87.51 (42.79–163.5) | 98.18 (19.43–208.2) | **0.949** |
| | **PGP9.5 MFI** | 6149 (4601–7283) | 5003 (3449–5804) | 0.010 | 4878 (3611–5855) | 5162 (2273–5802) | **0.455** |
| | **TrkA MFI** | 727 (474–1105) | 4021 (2227–5534) | **< 0.0001** | 4143 (2250–5552) | 3883 (1228–5706) | **0.881** |
| | **Norm TrkA MFI** | 0.13 (0.08–0.18) | 0.93 (0.56–1.25) | **< 0.0001** | 0.83 (0.6–1.22) | 0.98 (0.53–1.28) | **0.639** |
| Sub | **PGP9.5 area** | 562.9 (261.7–928.2) | 687.4 (390.2–999.0) | **0.441** | 709.3 (436.2–1024) | 552.5 (196.9–876.0) | 0.225 |
| | **PGP9.5 MFI** | 7728 (6172–10588) | 4829 (3481–6006) | **< 0.0001** | 4915 (3951–5564) | 3084 (4332–6085) | **0.289** |
| | **TrkA MFI** | 684.2 (548.7–1312) | 3502 (1600–4772) | **< 0.0001** | 3781 (2315–4880) | 3209 (1219–4729) | 0.360 |
| | **Norm TrkA MFI** | 0.1 (0.06–0.14) | 0.77 (0.5–0.92) | **<0.0001** | 0.8 (0.52–0.94) | 0.68 (0.42–0.94) | 0.416 |

Data are given as median and ranges. Abbreviations: Epi = epidermis region; Sub = sub-epidermis region; Norm = normalized to PGP9.5 intensity (TrkA MFI/ PGP9.5 MFI); MFI = mean fluorescence intensity.

### eTable_3

**GAP43/PGP9.5/CGRP Immunofluorescence Intensity Quantification**

| | Nerve type | Parameters | Control | All BIPN | *p* Value | Painless BIPN | Painful BIPN | *p* Value |
|---|---|---|---|---|---|---|---|---|
| **Epi** | **Peptidergic** | **GAP43 MFI** | 3479 (2221–5165) | 3366 (1679–4750) | 0.670 | 3001 (1659–4625) | 3946 (2452–5756) | 0.324 |
| | | **Norm GAP43 MFI** | 1.96 (0.1–3.41) | 1.2 (0.81–2.76) | 0.263 | 1.2 (0.81–2.74) | 1.63 (0.67–3.45) | > 0.999 |
| | **All** | **GAP43 MFI** | 2364 (1416–3145) | 2568 (2040–3524) | 0.141 | 2827 (2273 – 3570) | 2192 (1248 – 3311) | 0.090 |
| | | **Norm GAP43 MFI** | 1.32 (0.79–1.71) | 1.37 (1.1–1.71) | 0.558 | 1.37 (1.16–1.73) | 1.32 (0.62–1.56) | 0.137 |
| **Sub** | **Peptidergic** | **GAP43 MFI** | 3572 (2026–5317) | 2920 (2103–3738) | 0.297 | 3059 (2275–3960) | 2460 (1666–3613) | 0.233 |
| | | **Norm GAP43 MFI** | 1.18 (0.64–2.09) | 1.16 (0.75–1.44) | 0.678 | 1.19 (0.76–1.44) | 0.90 (0.61–1.68) | 0.633 |
| | **All** | **GAP43 MFI** | 2656 (1543–4207) | 2692 (1763–3313) | 0.510 | 2844 (2146–3411) | 1819 (1265–3019) | 0.041 |
| | | **Norm GAP43 MFI** | 1.32 (0.79–1.65) | 1.38 (0.9–1.67) | 0.841 | 1.41 (1.09–1.67) | 1.06 (0.81–1.69) | 0.156 |

Data are given as median and ranges. Abbreviations: Epi = epidermis region; Sub = sub-epidermis region; Norm = normalized to PGP9.5 intensity (TrkA MFI/ PGP9.5 MFI); MFI = mean fluorescence intensity.

### eTable_4

**Pain Assessment in Painful Patients using Numeric Rating Scale (NRS)**

| ID | Analgesic treatment | | Current pain | | | Maximum pain last week | | |
|---|---|---|---|---|---|---|---|---|
| | Yes/No | NRS reduction | NRS | NRS_corrected | Pain levels | NRS | NRS_corrected | Pain levels |
| 001 | Yes | 3 | 0 | 3 | Mild | 3 | 6 | Moderate |
| 002 | No | 0 | 0 | 0 | No | 8 | 8 | Severe |
| 003 | No | 0 | 5 | 5 | Moderate | 10 | 10 | Severe |
| 004 | Yes | 1 | 2 | 3 | Mild | 3 | 4 | Moderate |
| 005 | No | 0 | 0 | 0 | No | 8 | 8 | Severe |
| 006 | No | 0 | 0 | 0 | No | 3 | 3 | Mild |
| 007 | No | 0 | 0 | 0 | No | 3 | 3 | Mild |
| 008 | No | 0 | 7 | 7 | Severe | 6 | 6 | Moderate |
| 009 | No | 0 | 3 | 3 | Mild | 8 | 8 | Severe |
| 010 | No | 0 | 0 | 0 | No | 6 | 6 | Moderate |
| 011 | No | 0 | 2 | 2 | Mild | 8 | 8 | Severe |
| 012 | Yes | 5 | 3 | 8 | Severe | 3 | 8 | Severe |
| 013 | Yes | 1 | 1 | 2 | Mild | 3 | 4 | Moderate |
| 014 | No | 0 | 0 | 0 | No | 9 | 9 | Severe |
| 015 | Yes | 4 | 0 | 4 | Moderate | 8 | 12 | Severe |
| 016 | Yes | 2 | 2 | 4 | Moderate | 6 | 8 | Severe |
| 017 | Yes | 3 | 0 | 3 | Mild | 5 | 8 | Severe |
| 018 | No | 0 | 0 | 0 | No | 5 | 5 | Moderate |
| 019 | Yes | 3 | 0 | 3 | Mild | 5 | 8 | Severe |

**eFigure_1**

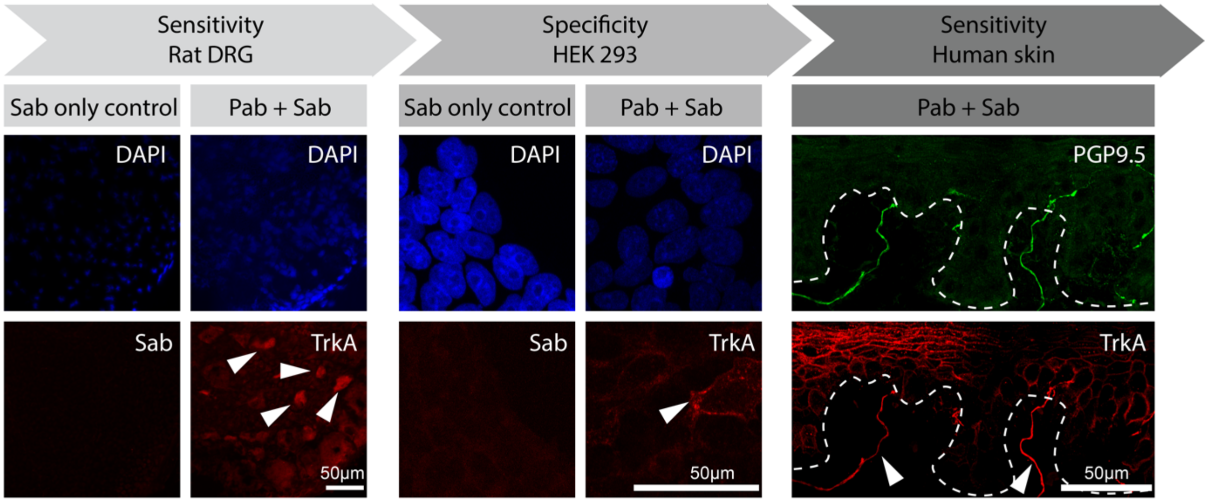


**Validation of TrkA Antibody Specificity and Sensitivity**

Left panel: Sensitivity testing in rat dorsal root ganglion (DRG) tissue. A secondary antibody-only control (Sab, first column) demonstrates minimal non-specific binding. Immunoreactivity of small DRG neurons is shown with the TrkA primary antibody (second column). Middle panel: A secondary antibody-only control (Sab, first column) demonstrates minimal non-specific binding in human-TrkA-transfected HEK293 cells. Immunoreactivity of human-TrkA-transfected HEK293 cells is shown with the TrkA primary antibody (second column). Right panel: Human skin staining confirming TrkA sensitivity. Scale bar = 50 µm. Abbreviations: Sab = secondary antibody; Pab = primary antibody.

**eFigure_2**

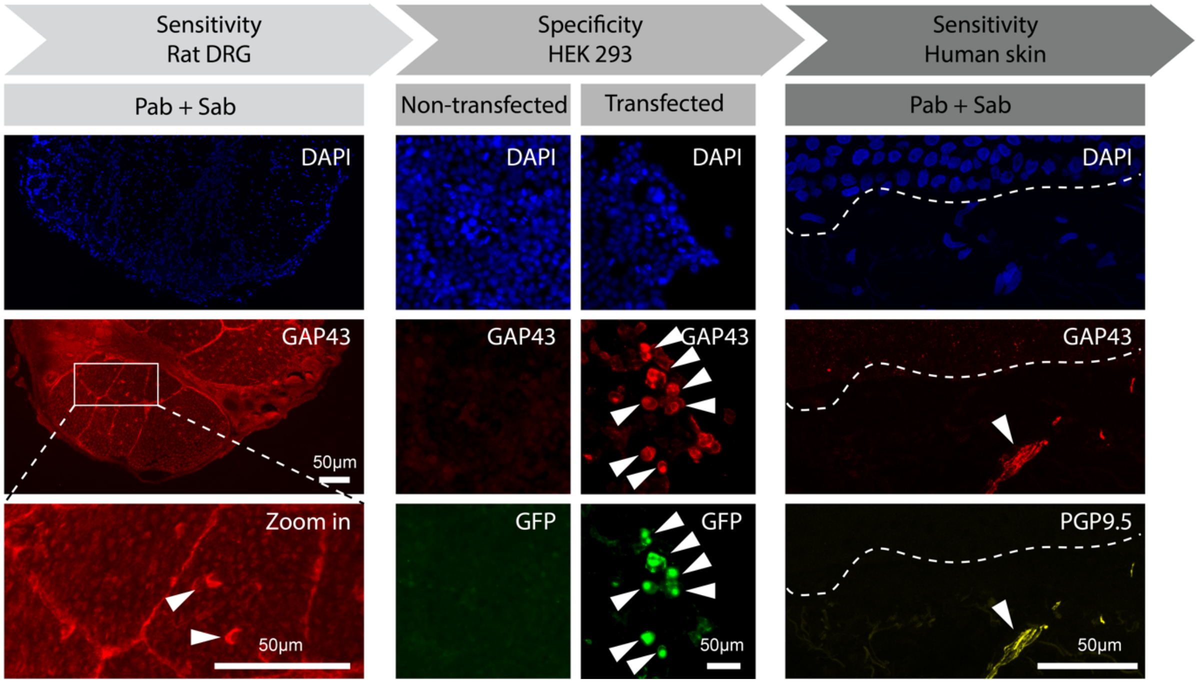


**Validation of GAP43 Antibody Specificity and Sensitivity**

Left panel: Sensitivity testing in rat dorsal root ganglion (DRG) tissue using the GAP43 antibody. Middle panel: Specificity testing using non-transfected (first column) and human-GAP43-transfected HEK293 cells (second column) with the GAP43 antibody. Right panel: Human skin staining confirming GAP43 sensitivity. Scale bar = 50 μm. Abbreviations: Sab = secondary antibody; Pab = primary antibody.

### eFigure_3

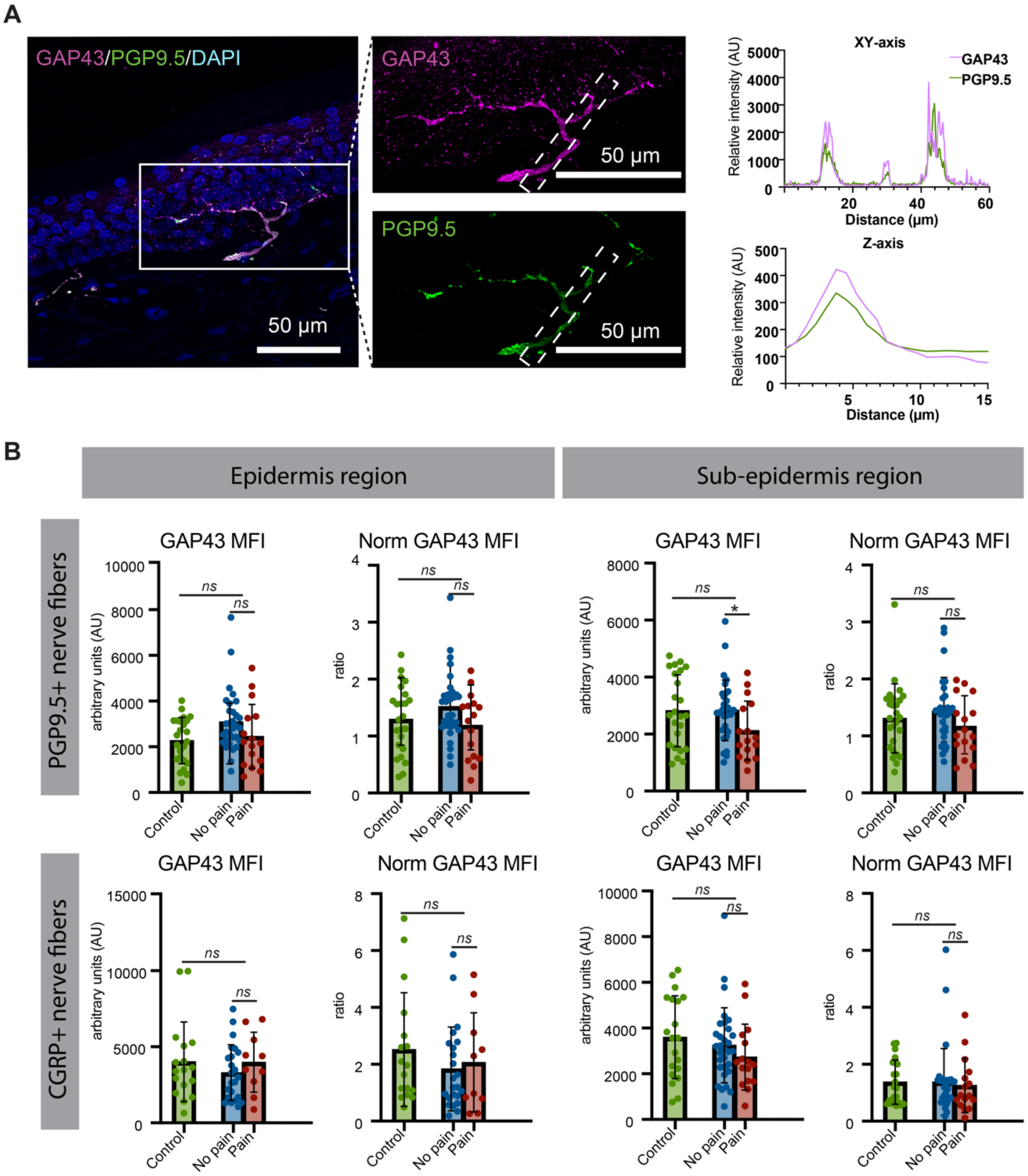


**GAP43 Immunofluorescence in Different Types on Nerve Fibers in Human Skin**

(A) Left: Representative immunofluorescence images showing GAP43 immunofluorescence, with PGP9.5 used as a reference marker for nerve fibers. Right: Colocalization plot for GAP43 and PGP9.5. (B) Mean fluorescence intensity of GAP43 and normalized GAP43 mean fluorescence intensity on whole nerve fibers and peptidergic nerve fibers in the epidermis region as well as sub-epidermis region.

Statistical analysis was performed using the Mann-Whitney U test for comparisons between control and patient groups, as well as between pain and painless subgroups. Scale bar = 50 μm. Abbreviations: MFI = mean fluorescence intensity; Norm = normalized to corresponding nerve fibers. * $p < 0.05$, ** $p < 0.01$, *** $p < 0.001$, ns: not significant.

**eFigure_4**

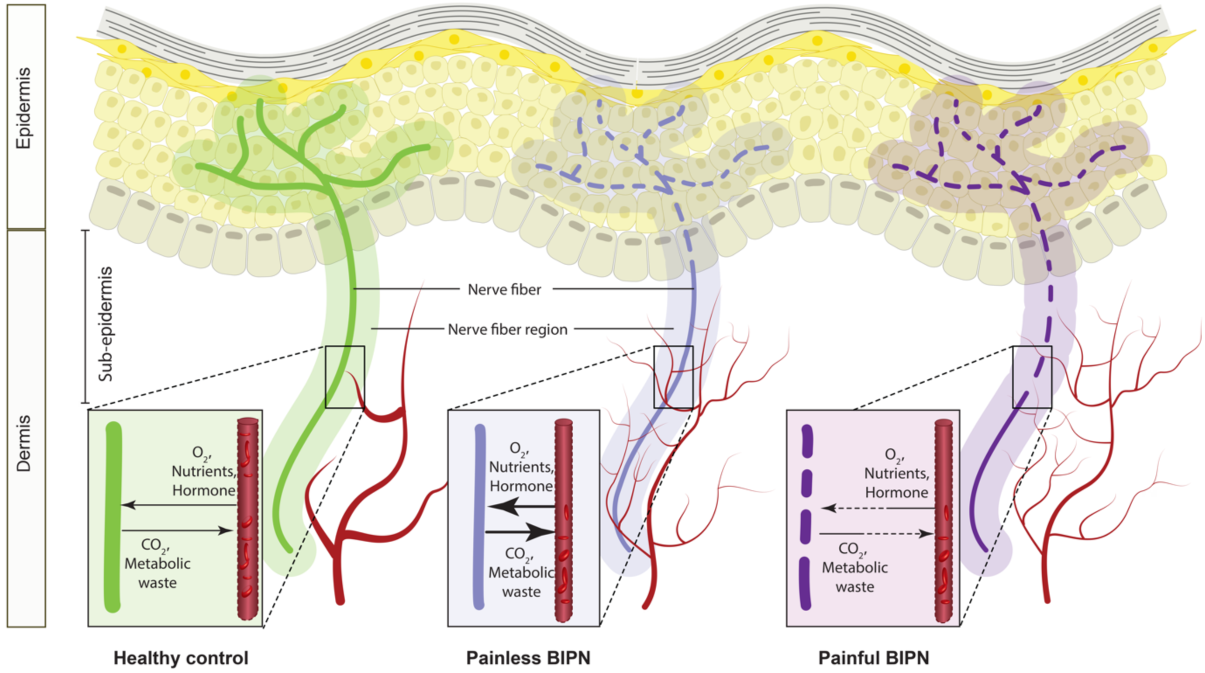


**Vascular-nerve Fiber Interaction in Human Skin**

Nerve Fiber Region: A 5 µm radius surrounding each nerve fiber was designated to examine close associations with blood vessels. Sub-epidermis Region: A 50 µm-deep area below the basement membrane was defined to focus on interactions in the superficial dermis.

**eFigure_5**

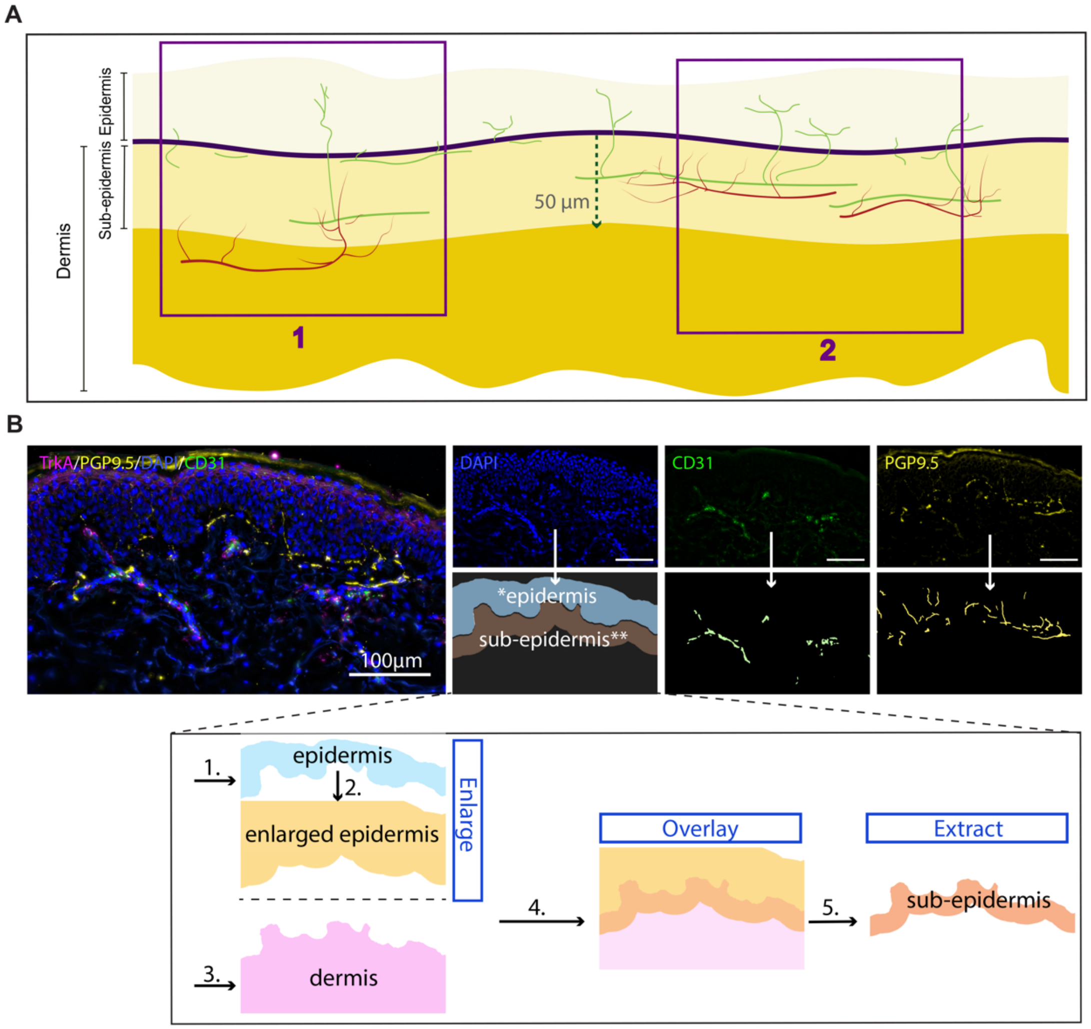


ROI selection and segmentation strategy

A: Skin biopsy cross-section with ROIs (purple) over epidermis, nerves (green), blood vessels (red), and 50-µm dermal region beneath basement membrane (dark blue). B: Deep-learning segmentation identifies nerves (yellow), vessels (green), epidermis (blue), and subepidermis (orange, extracted by computationally expanding the epidermis and overlapping with the dermis, strategy shown in the panel below). Scale bar = 100 µm.
